# Altered gene expression associated with postoperative delirium in patients undergoing surgery and anesthesia

**DOI:** 10.1101/2025.05.20.25320151

**Authors:** Maria Heinrich, Anna Fournier, Anna-Rosa Krüger, Florian Lammers-Lietz, Roland Krause, Peter Nürnberg, Reinhard Schneider, Georg Winterer, Maik Pietzner, Claudia Spies

**Affiliations:** Charité – Universitätsmedizin Berlin, corporate member of Freie Universität Berlin and Humboldt Universität zu Berlin, Department of Anesthesiology and Intensive Care Medicine, Augustenburger Platz 1, 13353 Berlin; Berlin Institute of Health at Charité (BIH) – Universitätsmedizin Berlin, BIH Academy, Clinician Scientist Program, Charitéplatz 1, 10117 Berlin, Germany; Bioinformatics Core, Luxembourg Center for Systems Biomedicine (LCSB), University of Luxembourg, Belvaux, Luxembourg; Swiss Data Science Center (SDSC), ETH Zurich and EPFL, Zurich, Switzerland; Cologne Center for Genomics, University of Cologne, Cologne, Germany Institute for Genetics of the University of Cologne, Cologne, Germany Atlas Biolabs GmbH, Berlin, Germany; PI Health Solutions GmbH, Berlin, Germany; Pharmaimage Biomarker Solutions Inc., Cambridge MA, USA; Computational Medicine, Berlin Institute of Health at Charité – Universitätsmedizin Berlin, Berlin, Germany; Precision Healthcare University Research Institute, Queen Mary University of London, London, UK

**Keywords:** gene expression, transcription, mRNA, immune system, (postoperative) delirium, surgery, anesthesia

## Abstract

Postoperative delirium is a severe complication associated with poor overall and especially neurocognitive prognosis after anesthesia and surgery. As a systemic phenomenon, peripheral immune response to surgical trauma may play a central role. Although analysis of differential gene expression in peripheral immune cells could provide insights into immune dysregulation in postoperative delirium (POD), no such analysis has been conducted yet in a sufficiently sized prospective cohort.

We performed gene expression analysis in N=599 cognitively healthy male and female patients ≥65 years who provided blood samples for microarray-based transcriptomics before major elective surgery and on the first postoperative day. Patients were followed up for delirium until the seventh postoperative day. We identified differentially expressed genes in POD using a multivariable linear regression framework adjusted for sex, age, body mass index, preoperative physical status, duration of anesthesia and operative procedure.

Preoperative gene expression was not significantly different in patients who were later diagnosed with POD. However, we identified a total of 1063 unique significantly associated genes which differed in baseline-corrected mRNA abundance among POD patients after surgery (n=394 positively, n=681 inversely). This set was significantly enriched for genes related to cellular and humoral immune response, RNA metabolism and platelet function.

Post-, but not preoperative gene expression in peripheral immune cells has been found to be altered in patients with POD. Whereas most enriched pathways were related to immune response and acute phase reaction, few molecular alterations were found, which may reflect nervous system alterations and need further clarification.

**HIGHLIGHTS:**

- Postoperative delirium (POD) is a common severe complication in older surgical patients
- Systemic inflammation has been considered a major hallmark of POD
- We describe immune cell gene expression in a large prospective cohort of surgical patients
- POD is associated with postoperative, but no preoperative alteration in gene expression
- Differentially expressed genes are involved in immune, platelet, but also neuronal function

## 1 Introduction

Delirium is defined by disturbances in attention, awareness, cognition, psychomotor behavior and emotional state. According to the 5^th^ edition of the Diagnostic and Statistical Manual of Mental Disorders (American Psychiatric Association), delirium is a consequence of another medical condition, intoxication or withdrawal, or multietiological. Delirium incidence after anesthesia and surgery (postoperative delirium, POD) ranges from 5% to 50% for different patient cohorts and surgical procedures (1). POD incidence is assumed to rise with increasing life expectancy, i.e., for Europe, the World Health Organization has predicted life expectancy of 80y in 2050 compared to 60y in 1950 (2). The relevance of POD for aging societies is further precipitated by the fact that surgical procedures are more often necessary in the most vulnerable group of older adult patients (3).

Patients who experienced POD have long-lasting impairment of physical functioning (4, 5). POD is a distressing experience for patients associated with posttraumatic stress disorder and lasting reduction of quality of life (6, 7). Neurocognitive disorders (NCD) may persist after POD and is associated with early exit from paid employment, social transfer payment dependence, increased care dependency, and mortality (8–10).

There are currently only symptomatic measures to prevent or treat POD due to a poor understanding of its molecular mechanisms (11–16). However, inflammation has long been considered a major pathomechanism in POD (17–19). Recently, Dillon and Vasunilashorn et al. analysed cerebrospinal fluid (CSF) samples from 24 patients experiencing POD after orthopedic surgery in spinal anesthesia as well as 24 matched controls (20). The identified POD-associated CSF proteins were involved in inflammatory response pathways, immune cell movement, function and turnover. This study provided novel insights into the role of neuroinflammation in POD, but the role of peripheral immune response for a neuroinflammatory spill-over is still under debate. To date, there have been reports on higher neutrophil-to-lymphocyte ratio in non-surgical patients with delirium (21). More indepth analyses have also suggested the involvement of particular immune cell subsets (22) and peripheral immune mediators, i.e., IL6 and TNFα (23, 24) as well as IL8, which acts as a chemoattractant for (25). It has been long proposed that senescence-associated alterations of CNS immune cells may predispose for delirium in presence of a sufficient peripheral inflammatory trigger (17). However, some researchers have reported that patterns of peripheral immune and neuroinflammatory response may be similar (26–28) and hence, it appears plausible that also immune dysregulation at the peripheral level may trigger neuroinflammation and POD. E.g., IL6 and TNFα, which are well-known mediators of sickness behavior) may link peripheral immune response and delirium (23, 24).

Within this context, gene expression analysis from peripheral immune cells may provide dual benefit for the understanding of POD pathogenesis. Firstly, transcriptome analysis can provide valuable information about POD-related processes at a molecular level, targeting a broad spectrum of potentially altered pathways. Secondly, it addresses changes of the peripheral immune system in POD, elucidating the role of systemic rather than neuroinflammation for POD. One gene methylation study and two previous postoperative gene expression studies in delirium have yielded promising results (29–31). However, the strength of the evidence remains low due to limited sample size, case-control designs and insufficient information about gene expression patterns before delirium occurrence as well as postoperative changes. Hence, gene expression data from large prospective cohorts of patients at risk for POD are still missing.

Here, we present the so far largest study (BioCog) on changes in peripheral immune cell gene expression possibly predating or accompanying POD in 599 surgical patients. We describe substantial changes in gene expression among POD patients following surgery that were not observable before surgery indicating systemic effects.

## 2 Methods

### 2.1 Study design and participants

The BioCog (Biomarker Development for Postoperative Cognitive Impairment in the Elderly, www.biocog.eu) study is an EU-funded multicenter prospective observational cohort study with the aim of identifying risk factors for POD.

Female and male patients were enrolled in two tertiary care centers at the Charité– Universitätsmedizin Berlin, Germany, and the University Medical Center Utrecht, Netherlands. Consenting patients aged ≥65 years presenting for elective non-cardiac surgery with an expected duration >60min were included. Patients were excluded from the study according to the following criteria:

- positive screening for pre-existing major neurocognitive disorder defined as a MiniMental Status Examination (MMSE) score ≤23 points
- any condition interfering with neurocognitive assessment (severe sensory impairment, neuropsychiatric illness including alcohol and drug dependence, intracranial surgery)
- unavailability for follow-up assessment
- accommodation in an institution due to official or judicial order
- inability to give informed consent (including detection of preoperative delirium assessed at baseline)

The study was conducted in line with the declaration of Helsinki (clinicaltrials.gov: NCT02265263). All procedures were approved by the local ethics committees in Berlin, Germany (EA2/092/14) and Utrecht, Netherlands (14–469). All participants gave written informed consent prior to inclusion.

Sex, but not gender, was recorded as either male and female, without a standard operating procedure for the assessment of sex. That is, research staff was allowed to record sex according to both the patient’s self-report or the patient file.

### 2.2 Sample size calculation

The major aim of BioCog was the development of a POD prediction algorithm and the sample size (N=1200 patients) was planned a priori according to this purpose. No sample size calculation was performed with regard to gene expression analysis.

### 2.3 Delirium screening

POD during the first seven days after surgery was the primary endpoint. Independently of the routine hospital procedures, POD screening was started in the post-anesthesia recovery room and repeated twice per day at 8:00am and 7:00pm (±1h) up to seven days after surgery. All study visits were made by study physicians or trained study nurses and study assistants under supervision of a study physician. POD was defined according to DSM-5 criteria and supplemented by prospective screening with four validated tools which were recorded at each visit to improve sensitivity (12, 32). Patients were considered delirious if at least one of the following criteria was positive:

- ≥2 cumulative points on the Nursing Delirium Screening Scale (Nu-DESC) (33), and/or
- a positive Confusion Assessment Method (CAM) score on a general ward (34), and/or
- a positive CAM for the Intensive Care Unit (CAM-ICU) score on an ICU (35), and/or
- patient’s chart review showing descriptions of delirium (e.g. confused, agitated, drowsy, disorientated, delirious, received antipsychotic therapy)

The researchers involved in the analyses presented here were not blinded regarding the outcome.

### 2.4 Transcriptomics

Blood was collected before anesthesia, on the first postoperative day and after 3 months (day +90) in PAXgene blood RNA tubes (Qiagen, Hilden, Germany). For this study, only microarray-based RNA data until the first postoperative day have been analyzed.

Total RNA, including RNA longer than approximately 18 nucleotides, was isolated by means of the PAXgene blood RNA isolation kit (Qiagen, Hilden, Germany) according to the manufacturer’s instructions. Total RNA (RIN > 6.0) was processed and hybridized to microarrays according to Affymetrix’ specifications. RNA amplification and microarray preparation were done by ATLAS Biolabs GmbH (Berlin, Germany). 100ng of mRNA per fraction was amplified and loaded onto Affymetrix Clariom S human microarray plate for 96 samples (Thermo Fischer, Santa Clara, CA, USA) in accordance with manufacturer’s recommendations. Hybridization, washing, staining and imaging took place in the GeneTitan™ Multi-Channel (MC) Instrument to provide the automated array processing. Both spike control oligos and hybridization control stages of the procedure were performed according to the manufacturer’s instructions and under quality control. Raw data were normalized with the robust multiarray average method implemented in the Affymetrix Expression Console software (Thermo Fischer, Santa Clara, CA, USA). Further, all data were quality checked with R-package arrayQualityMetrics to assess the reproducibility, identify apparent outlier arrays and noise (36).

### 2.5 Statistical analysis

We analysed differences in gene expression between patients with and without POD using multivariable linear regression models, treating preoperative and postoperative transcript abundance as the dependent variable and POD as the independent variable of interest. Regression models of preoperative gene expression were adjusted for sex, age, body mass index, American Society of Anesthesiologists (ASA) physical status (≥III) as a surrogate parameter for preoperative morbidity, duration of anesthesia and operative procedure (eq. 1). Regression models of postoperative transcript abundance included the same independent variables and were also adjusted for preoperative transcript abundance (eq. 2):

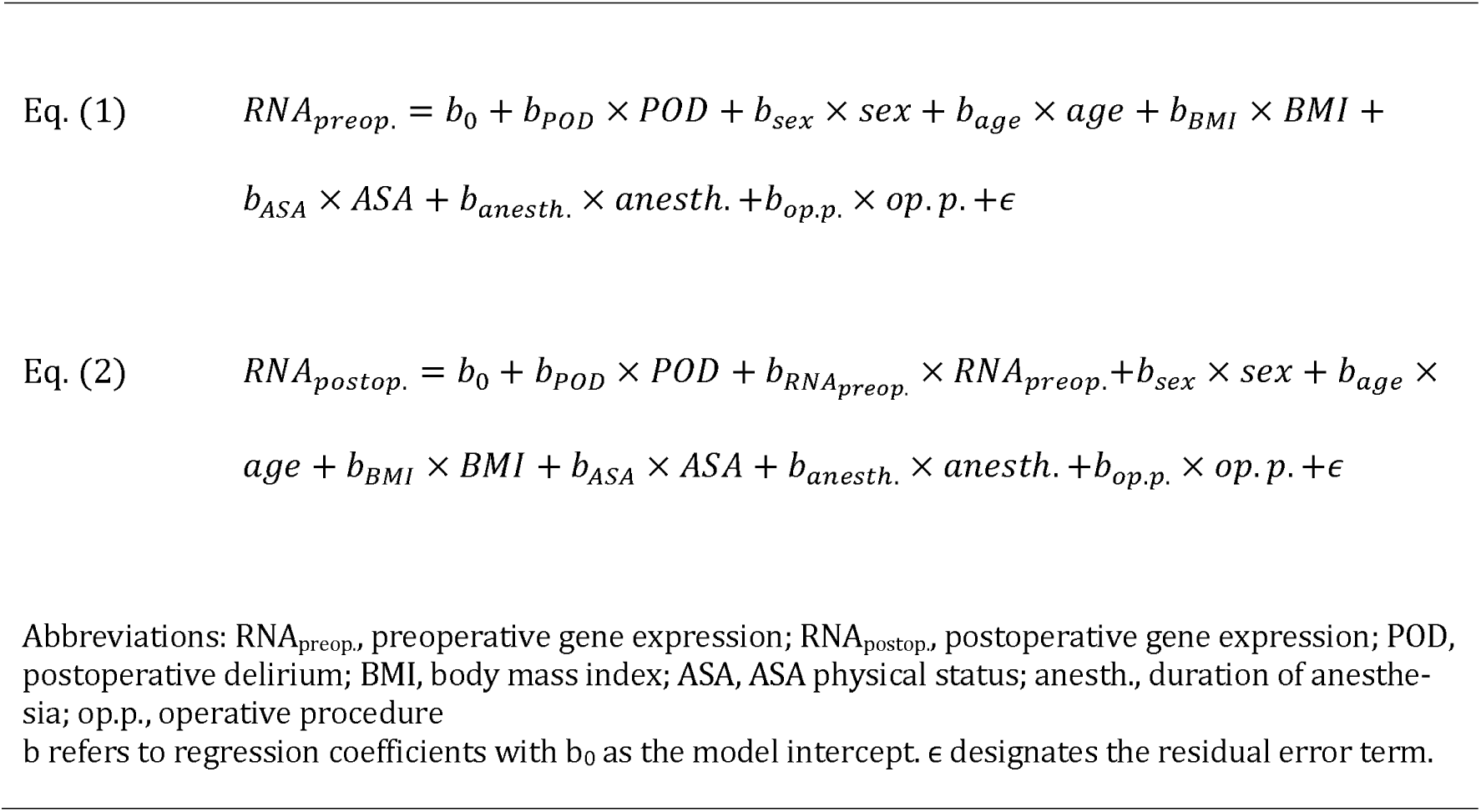

For both regression analyses of pre- and postoperative gene expression, we report log-fold change and p-values for POD occurrence. The level of significance was set to p≤0.05 after Benjami-Hochberg correction for multiple testing. R software version 4.1.2. was used for all statistical analyses.

### 2.6 Gene enrichment analysis

We used g:Profiler for R software (gprofiler2, v. 0.2.1) for pathway and entity analysis using the gene ontology (GO), Kyoto Encyclopedia of Genes and Genomes (KEGG) and Reactome (REAC) databases. The Fisher test implemented in the GOSt function in gprofiler2 for R software was used to appraise over- and under-representation in the pathways and biological entities. The level of significance was set to p≤0.05 using Benjamini-Hochberg correction for multiple testing.

No imputations of missing data was performed (complete case analysis) and no outliers have been removed before analysis. Sex was considered as a relevant confounder (37–40) in all analyses, but with respect to sample size considerations and multiple testing, we do not present sex-specific results. Transcripts of gonosomal genes have not been considered in this work.

## 3 Results

### 3.1 Description of the cohort

In total, 599 patients with a mean age of 73±5 years and a preoperative MMSE of 28.6±1.4 points were included in the analysis. Mean body mass index (BMI) was 27.3±4.9kg/m². 246 (41%) of the sample were women, and 325 (54%) underwent peripheral surgery. Mean duration of surgery was 146.9±117.8 minutes (min), and duration of anesthesia was 249.5±203.2min in the whole sample. Of 599 patients, 126 (21%) developed postoperative delirium. Figure 1 displays the patient flow chart and table describes the cohort. Overall, 20896 transcripts were analyzed.

**Figure 1:**
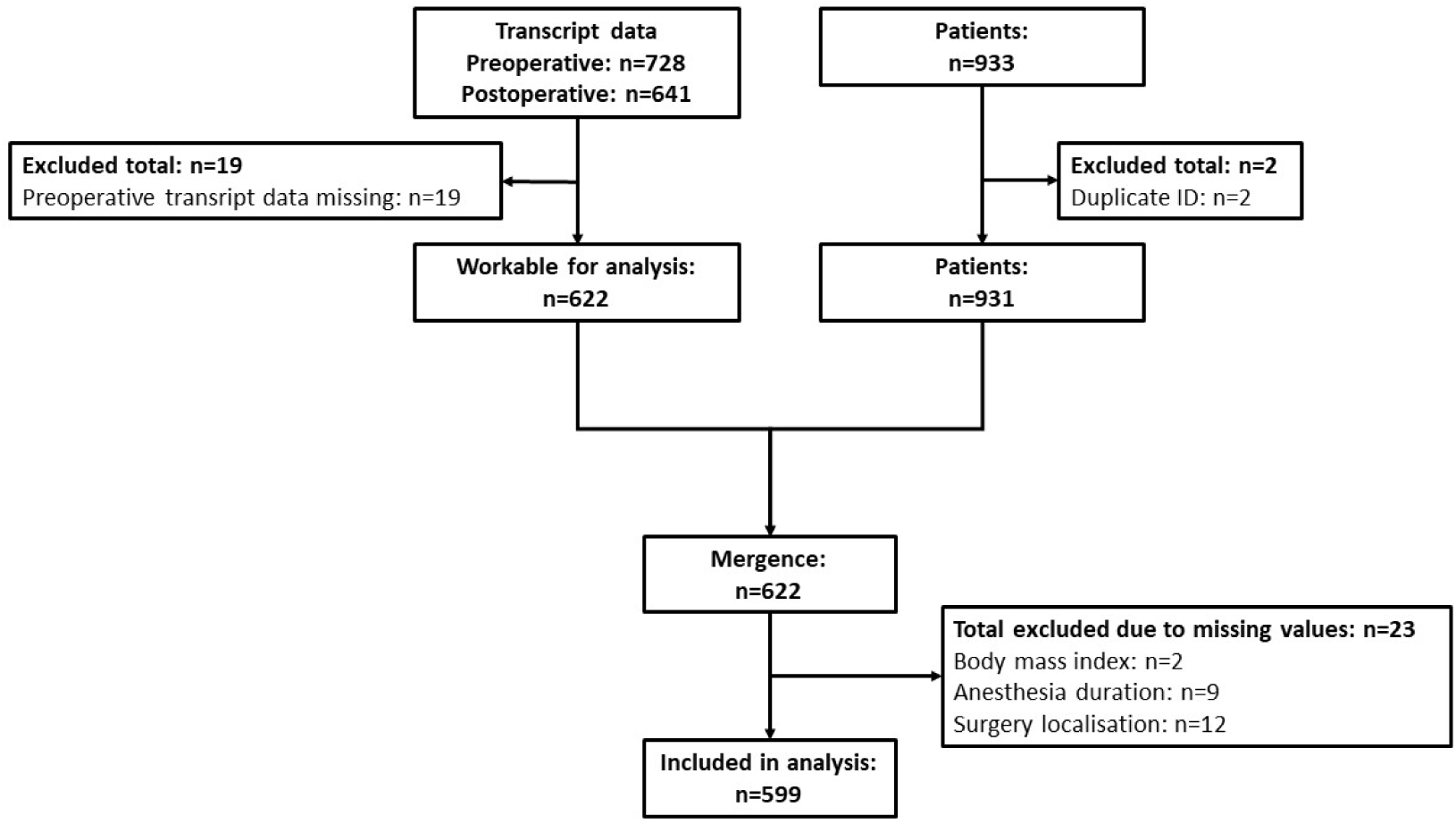
Patient flow chart

### 3.2 Results of mRNA analyses

#### 3.2.1 Preoperative alterations in mRNA abundance related to POD

We did not observe statistically significant differences passing multiple testing correction comparing gene signatures between POD and non-POD patients before surgery after adjustment for sex, age, body mass index, ASA physical status, duration of anesthesia and operative procedure, suggesting no differences in preoperative transcript abundance.

#### 3.2.2 Postoperative alterations in mRNA abundance related to POD

We found that 1075 transcripts of 1063 unique genes had altered postoperative expression leves among POD patients after adjustment for preoperative gene expression, sex, age, body mass index, ASA physical status, duration of anesthesia and operative procedure (n=394 positively, n=681 inversely). Intermediate filament family orphan 2 (*IFFO2*), diphosphoinositol pentakisphosphate kinase 1 (*PPIP5K1*), caspase 7 (*CASP7*), ring finger protein 125 (*RNF125*), dachshund family transcription factor 1 (*DACH1*) and 15-hydroxyprostaglandin dehydrogenase (**HPGD**) were the most significantly associated genes (q<10^-2.2^), see figure 2).

**Figure 2:**
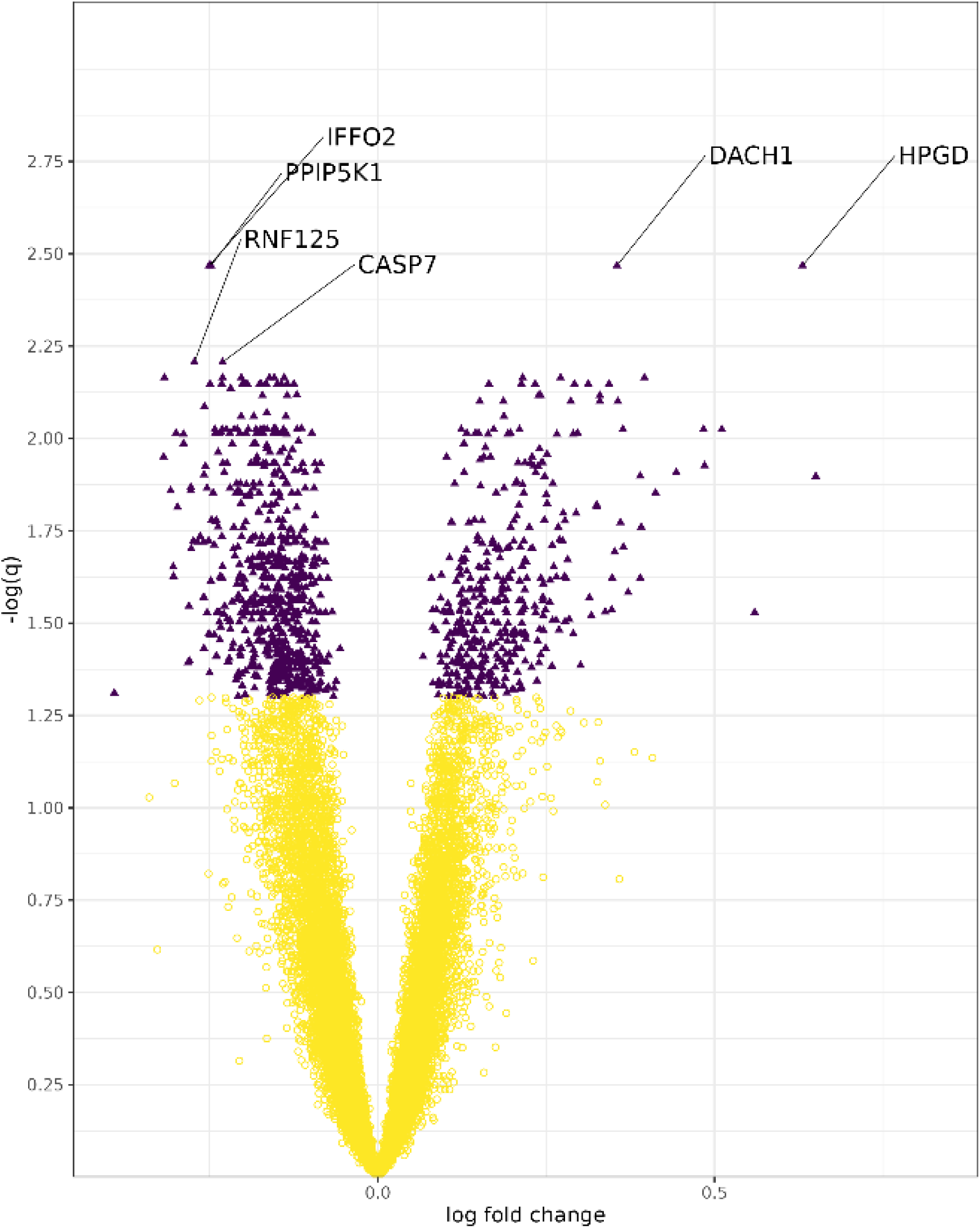
Volcano plot on postoperative differences in mRNA abundance between patients with and without POD. On the x-axis, the regression coefficient of a multiple regression model of postoperative transcript abundance on POD is given after adjustment for preoperative transcript abundance, sex, age, body mass index, ASA physical status (≥III), duration of anesthesia and operative procedure. Transcripts with a q-value < 0.05 are highlighted in purple, and transcripts with -log_q_>2.2 are tagged with the corresponding gene name. 19 transcripts were strongly lower in POD patients after surgery (log-fold change <-0.275): AK5, IFI44L, DOCK7, GBP5, TGFBR3, GNLY, STAT4, NR1D2, CCR3, FGFBP2, GZMA, FAM169A, IFIT3, IFIT1, KLRG1, KLRF1, EPSTI1 and CLC. Extreme postoperative upregulation was noted for four transcripts (log-fold change>0.5): HPGD, TENM1, OLAH and CD177.

#### 3.2.3 Pathway analysis of postoperative differences in mRNA abundance

Of 1063 unique genes which were found to be lower or higher expressed after surgery in patients with POD compared to patients without POD, we observed twelve significantly enriched or depleted pathways (see table 2). Genes from nine pathways were overrepresented in patients with POD. Among these, five pathways were directly linked to immune function, inflammation and host defense against pathogens, two were related to regulation of transcription and further two identified pathways were involved in hemostasis. Two out of three pathways with gene underrepresentation were related to perception and transmission of sensory information.

**Table 1:** Patient characteristics for POD analysis (n =599). Characteristics are given as absolute and relative numbers, except for those marked , which are given as mean with standard deviation. A sex-specific description is given in supplementary table S1.

|  | POD | No POD |
| --- | --- | --- |
|  | n=126 | n=473 |
|  | (21.04%) | (78.96%) |
| Women | 60 (47.62%) | 186 (39.32%) |
| Peripheral surgery | 44 (34.92%) | 281 (59.41%) |
| Age (years)<sup>a</sup> | 74.13 (5.26) | 72.08 (4.98) |
| Duration of anesthesia (minutes)<sup>a</sup> | 374.75 (254.89) | 216.14 (172.70) |
| BMI (kg/m<sup>2</sup>)<sup>a</sup> | 27.68 (5.50) | 27.21 (4.48) |
| ASA physical status I-II | 57 (45.24%) | 331 (69.98%) |

**Table 2:**
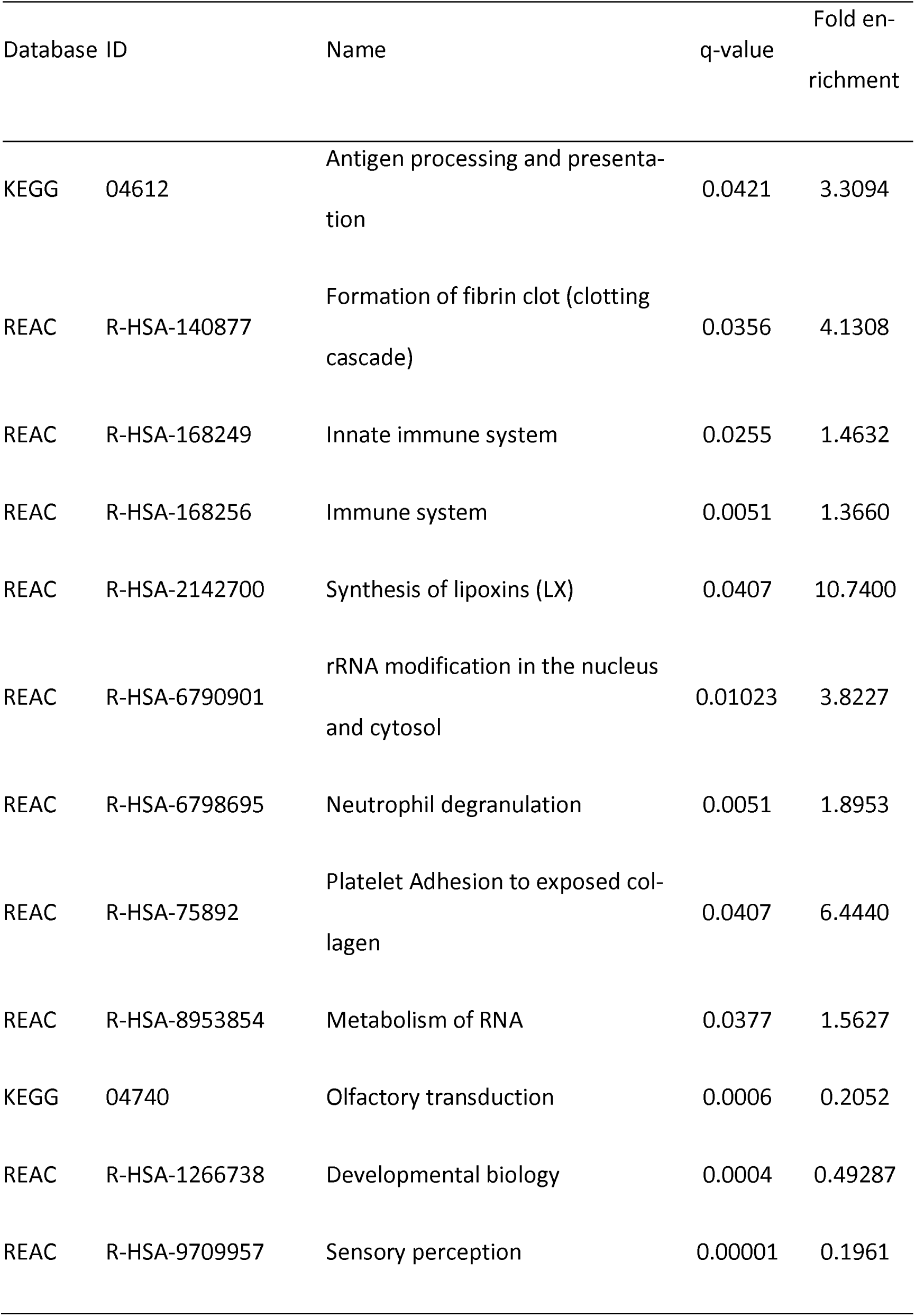
Overview on pathways altered in patients with POD after surgery and anesthesia

#### 3.2.4 Gene ontology analysis of postoperative differences in mRNA abundance

Table 3 lists the biological processes with the strongest association with POD from each ontology (biological processes, cellular components and molecular function). In general, the enriched ontologies were unspecific and located at high positions in the hierarchy. To describe more specific pathomechanisms in POD as possible targets for interventions, we present those ontologies which are over- or underrepresented, but do not have any offspring ontologies.

**Table 3:**
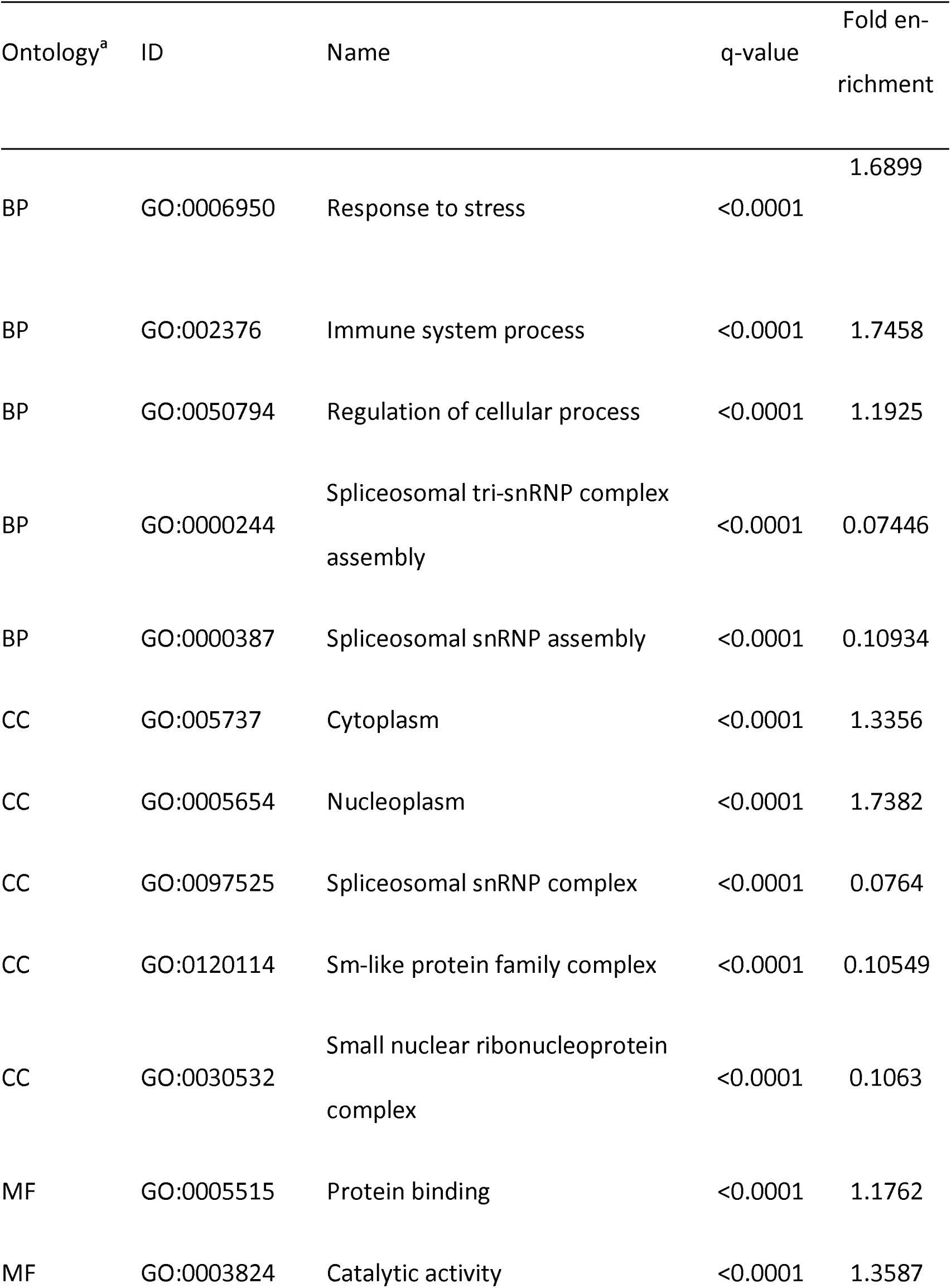
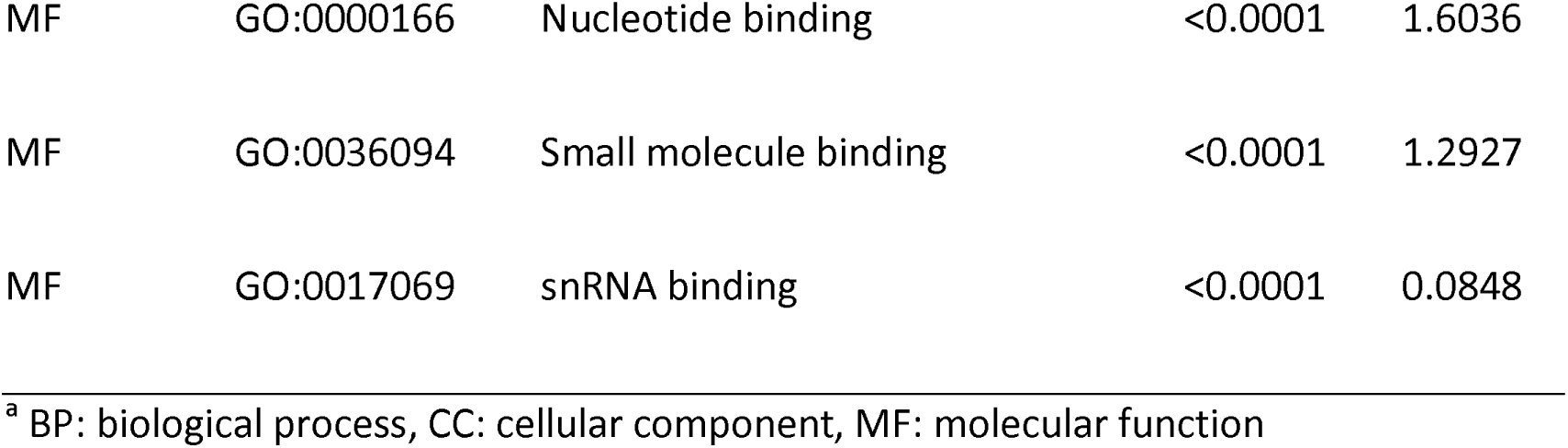
List of gene ontologies with the strongest association with POD. For reasons for clarity, the table is limited to the five ontologies with the strongest association with POD.

540 biological processes were over- and 29 underrepresented. Among these, we identified seven biological processes without further offspring ontologies which were all overrepresented (see table 4).

**Table 4:** List of biological process ontologies without offspring ontologies.

| ID | Name | q-value | Fold enrichment |
| --- | --- | --- | --- |
| GO:0006337 | Nucleosome disassembly | 0.0077 | 7.1183 |
| GO:0097011 | Cellular response to granulocyte macrophage CSF stimulus | 0.0221 | 8.1968 |
| GO:0042325 | Regulation of phosphorylation | 0.0257 | 1.5168 |
| GO:1903238 | Positive regulation of leukocyte tethering or rolling | 0.0341 | 6.9358 |
| GO:1903298 | Negative regulation of hypoxia-induced intrinsic apoptotic signaling pathway | 0.0344 | 9.6605 |
| GO:0048250 | Iron import into the mitochondrion | 0.0369 | 22.5413 |
| GO:1900099 | Negative regulation of plasma cell differentiation | 0.0369 | 22.5413 |

We identified 96 cellular complexes associated with POD, among which 10 were underrepresented. 14 overrepresented ontologies without offspring ontologies were identified (see table 5).

**Table 5:**
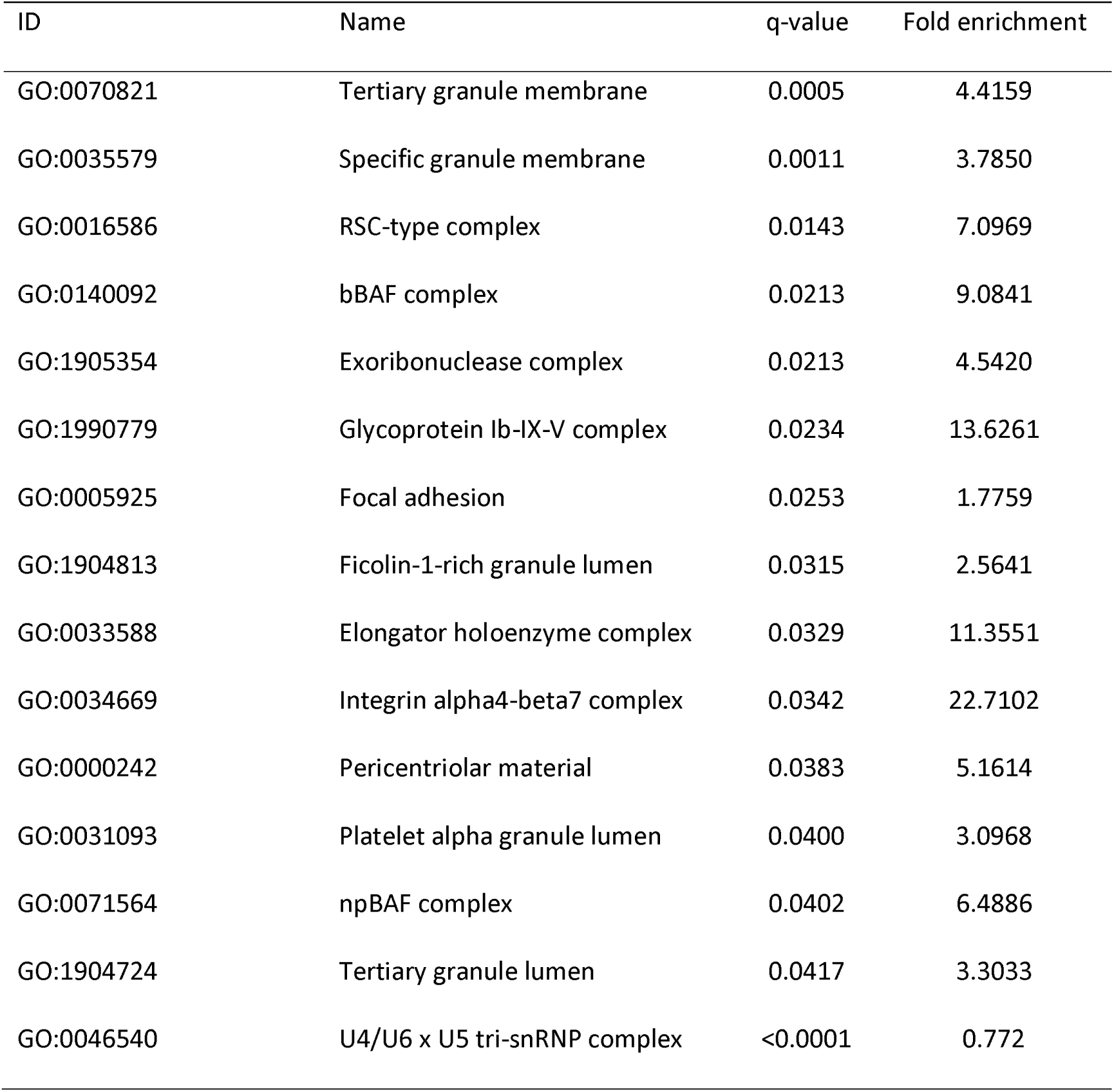
List of cellular complex ontologies without offspring ontologies.

Of 55 molecular functions, 49 were overrepresented. Two molecular functions without off-spring ontologies were identified (see table 6).

**Table 6:** List of molecular functions without offspring ontologies.

| ID | Name | q-value | Fold enrichment |
| --- | --- | --- | --- |
| GO:0003735 | Structural constituent of the ribosome | <0.0001 | 0.0922 |
| GO:0008607 | phosphorylase kinase regulator activity | 0.0102 | 20.7515 |

## 4 Discussion

Here, we present the so for largest prospective study of gene expression in peripheral immune cells to reveal potential pathomechanisms in POD (41).

Of note, differentially expressed genes related to POD were identified in postoperative blood samples only, whereas we observed no differences in preoperative samples. This is in line with our results from a gradient-boosted trees model for POD prediction (42): We observed that preoperative gene expression was a poor predictor of POD, but postoperative changes in transcript abundances could be employed for POD risk monitoring. It further emphasizes the role of surgical trauma and anaesthetic toxicity in the pathology of POD, and suggests that molecular dysregulations in POD are a pathological response to physical stress.

To the best of our knowledge there are only two independent previous studies investigating gene expression of peripheral immune cells in patients with delirium at all: The research team of Song et al. conducted microarray-based gene expression analyses in four delirious patients and four age-matched controls after orthopedic surgery (30). Both the results of Song and our work suggest a particular role for neutrophils, especially neutrophil degranulation and ficolin-1-rich granules. Ficolins were initially discovered as a transforming growth factor (TGF)-b1-binding protein, but current evidence indicates a role as innate soluble pattern recognition molecules which trigger the complement lectin pathway and mediate cellular detritus removal (43). In another case-control study with 51 patients, Kalantar et al. described complement system activation in delirium due to various etiologies by whole-genome mRNA expression analysis (29).

Beyond complement activation, it hast also been shown that ficolin-1 activates NF-kB signaling cascade to upregulate IL-8 production as a chemotactic signal for neutrophils (44). We and other researchers have previously reported an involvement of IL-8 in POD (25, 45–49). Various ontologies and pathways identified in our current gene expression analysis indicated a relevant role for immune, but especially neutrophil response in POD, e.g., neutrophil degranulation, with a particular role for ficolin-1 rich and tertiary granule functioning and their interaction with the SNARE complex (50, 51). Supporting our findings, Saito and collegues studied genome-wide DNA methylation patterns in another case-control study of delirium including 87 patients and provide evidence for involvement of pathways related to neutrophil activation, vesicle and tertiary granule function (31).

By transcriptomic profiling, Kalantar and colleagues found that alterations in mitochondrial structure and heme metabolism would differ between delirious and non-delirious patients (29). Mitochondrial iron uptake has also been identified as a potential dysregulated pathway in POD here. Previous works already described altered iron metabolism in a small sample of patients with delirium tremens (52) and an association of brain iron and incident delirium in an analysis of patients with hemochromatosis-associated mutations from the UK biobank cohort (53). In an exploratory analysis of magnetic resonance imaging data in the same cohort we observed an association between suspected iron-deposits in the central nervous cholinergic systems to be associated with POD and postoperative cognitive decline (54).

Notably, leukocyte tethering and rolling as well as associated integrin complexes have been identified in both our work, Saito (31) and the study by Kalantar (29). However, Kalantar described a relation of these pathways to infection and infection-related delirium rather than delirium itself. Especially integrin α4β7, which is relevant for leukocyte tethering and rolling, was identified as a potential pathomechanism in POD in our study. Integrin α4β7 may also play a role in multiple sclerosis, but relevance integrin -coding RNA expression in peripheral immune cells for neuroinflammation cannot be inferred from our data (55).

Lipoxins interact with various of the above-mentioned pathways, including neutrophil function, leukocyte trafficking, termination of the inflammatory response, macrophagic clearance of apoptotic detritus and gene transcription.

Various identified pathways and ontologies related to platelet function, especially α-granules. They contain von Willebrand factor, and hence link thrombus and fibrin formation. They contain a plethora of membrane-bound and soluble proteins, possibly mediating systemic effects beyond hemostasis (56), i.e., integrins, various receptors, and beyond platelet and coagulation activators, inflammatory mediators, antimicrobial substances, pro- and antiangiogenetic as well as mitogenetic factors in general.

Notably, although we investigated gene expression in peripheral immune cells, biomarkers of neurogenesis and neural tissue differentiation was altered in POD. The RSC (remodeling the structure of chromatin) complex, especially its components bBAF (brain-specific BRG1/BRM-associated factor) (57) and npBAF (neuronal progenitor BRG1/BRM-associated factor). Although the significance of neuronal biomarker alterations found in immune cells needs to be elucidated, similar observations have been reported by Song et al. as well as Saito et al. who described alterations in molecular pathways related to neurotransmission, and synapse formation (30, 31). The RSC complex plays a role in control of transcription and has essential functions in differentiation, e.g., of neuronal tissue, including adult neurogenesis and gliogenesis, impacting learning and memory formation (58). In addition, elongator holoenzyme complex was overrepresented in POD-related transcripts. Elongator controls transcription, especially during neurite formation and has been suggested to be involved in various neurological diseases, such as familial dysautonomia, amyotrophic lateral sclerosis and Rolandic epilepsy (59).

A major finding of gene expression analyses was the postoperative downregulation of the *CASP7* protease in patients with POD. By cleavage of key cell enzymes and reduction of toxic cell component release into the tissue, *CASP7* controls apoptosis and ameliorates the inflammatory reaction to dying cells (60). *CASP7* gene variants have already been associated with Alzheimer’s disease and mild cognitive impairment (61–63). It was also shown that *CASP7* is possibly involved in microglia activation, and inhibition of *CASP7* has neuroprotective effects by reducing microglial activation (64). Considering that microglial inflammation may be a major driver of POD, and that *CASP7* downregulation may be hence be expected to be neuroprotective, the interactions of apoptosis and neuroinflammation in POD warrant further investigation (65).

Apart from those studies cited above, we conducted a gradient-boosted trees model in the same data to assess the value of gene expression for POD prediction (42). Although we used the same dataset, only *HPGD* of the differentially expressed genes was identified as highly relevant for POD prediction, suggesting senescence as a pathomechanism in POD (66). However, gradient-boosted trees also identified transcripts involved in neuroinflammation (*BTN3A1*, (67)) and autophagy (*LAP3* (68))

### 4.1 Limitations

Enrichment analyses using the Gene Ontology database yielded a plethora of ontologies significantly associated with POD. However, these ontologies were located within the upper levels of the hierarchical tree and even vertically related, providing only little information about specific molecular mechanisms in POD. The result may be interpreted as holistic disturbance of cellular and molecular functions in POD, or as insufficient specificity of the analytical approach. Results may also be affected by POD due to various etiologies in our cohort which comprises a broad range of patients with diverse characteristics undergoing a heterogenous set of surgical procedures. Future studies may be advised to use alternative data reduction and clustering approaches to identify subgroups with certain RNA signatures among delirious patients to enlighten molecular mechanisms.

We observed multiple underrepresented entities in our analyses, including pathways and ontologies related to sensory perception, especially olfaction, and pre-mRNA splicing. Although the relevance of perception may be intuitive for POD, the interpretation of underrepresented pathways is difficult. As stated above, most over-expressed entities were unspecific, and hence, underrepresentation may be related to the choice of control genes in our analysis, rather than a pathomechanistic relation. In addition, patients with severe sensory impairment were excluded from the analysis, and hence, a relevant association between impaired perception and POD may be difficult to study in the BioCog sample.

Sex is a relevant confounder in studies on POD, since there is a well-documented, but poorly understood association of sex with POD risk (38–40). Furthermore, underlying pathomechanisms for POD may differ by sex (37). However, it is unknown to what extent the association of sex and POD may be genetically determined or rather be determined by environmental and behavioral factors such socioeconomic status and education, and health behavior. While we adjusted our gene expression analysis for sex, the sex-attributed POD risk may be partially mediated by sex-specific gene expression, and may hence be obscured by statistical adjustment. Instead, an adjustment for sex-related behavioral and environ-mental POD risk factors would be preferable to sufficiently address genetic and external mechanisms that lead to POD.

The transcriptomic analysis may provide new potential targets for novel therapeutic approaches for POD. However, our analysis does not provide information on the direction of dysregulation, the need for stimulating or inhibiting interventions, and further studies are needed before clinical trials can be designed.

### 4.2 Conclusion

Here, we present the so far largest prospective study of differential blood gene expression in POD. Although our results point to systemic transcriptional disturbances in POD, we identified several circumscribed molecular pathways warranting further research. Briefly, neutrophil function and leukocyte directing, iron metabolism, regulation of transcription in neuronal tissue and mediator release from platelets warrant further studies.

## 6 Author contributions

Maria Heinrich, MD: conceptualization, data curation, formal analysis, investigation, methodology, writing – original draft; Anna Fournier, PhD – methodology, formal analysis, writing – review and editing; Anna Rosa Krüger, BSc: data curation, formal analysis, investigation, methodology, visualization, writing – original draft; Roland Krause, PhD: data curation, methodology, writing – review & editing; Florian Lammers-Lietz, MD: data curation, investigation, writing - original draft; Peter Nürnberg, PhD: methodology, investigation; Reinhard Schneider, PhD: funding acquisition, conceptualization, resources; Georg Winterer, MD, PhD: conceptualization, funding acquisition, methodology, project administration; Maik Pietzner, PhD: methodology, supervision, writing – review & editing; Claudia Spies, MD, PhD: conceptualization, funding acquisition, methodology, project administration, resources, writing – review & editing

## Acknowledgements

We would like to express our gratitude to the whole BioCog study team, including the study team at the UMC Utrecht. PI Health Solutions GmbH (https://pihealthsolutions.com/) and the PharmaImage team are acknowledged for their extensive contributions during the conduct of the BioCog study.

## 8 Declaration of Generative AI and AI-assisted technologies in the writing process

At no stage of the writing process, artificial intelligence or the support of artificial intelligence was used for manuscript preparation.

## 9 Data availability

Participant data may be made available upon request following publication to researchers who provide a methodologically sound proposal in accordance with applicable legal and regulatory restrictions after careful review of each individual request. Access will only be granted in cases where the potential receiver of the data and purpose of the analysis is covered by the patients’ informed consent and applicable legal regulations.

Proposals for data analysis must be directed to both. Analyses will be limited to those approved in appropriate ethics and governance arrangements. All study documents which do not identify individuals (e.g. study protocol, informed consent form template) will be freely available on request.

## References

1. Fournier A, Krause R, Winterer G, Schneider R. Biomarkers of postoperative delirium and cognitive dysfunction. Front Aging Neurosci. 2015;7:112.

2. World report on Ageing and Health. Geneva, Switzerland: World Health Organisation; 2015 2015.

3. Fowler AJ, Abbott TEF, Prowle J, Pearse RM. Age of patients undergoing surgery. Br J Surg. 2019;106(8):1012–8.

4. Hshieh TT, Saczynski J, Gou RY, Marcantonio E, Jones RN, Schmitt E, et al. Trajectory of Functional Recovery After Postoperative Delirium in Elective Surgery. Ann Surg. 2017;265(4):647–53.

5. Kim DH, Afilalo J, Shi SM, Popma JJ, Khabbaz KR, Laham RJ, et al. Evaluation of Changes in Functional Status in the Year After Aortic Valve Replacement. JAMA Intern Med. 2019;179(3):383–91.

6. Drews T, Franck M, Radtke FM, Weiss B, Krampe H, Brockhaus WR, et al. Postoperative delirium is an independent risk factor for posttraumatic stress disorder in the elderly patient: a prospective observational study. Eur J Anaesthesiol. 2015;32(3):147–51.

7. Abelha FJ, Luis C, Veiga D, Parente D, Fernandes V, Santos P, et al. Outcome and quality of life in patients with postoperative delirium during an ICU stay following major surgery. Crit Care. 2013;17(5):R257.

8. Rudolph JL, Marcantonio ER, Culley DJ, Silverstein JH, Rasmussen LS, Crosby GJ, et al. Delirium is associated with early postoperative cognitive dysfunction. Anaesthesia. 2008;63(9):941–7.

9. Inouye SK, Marcantonio ER, Kosar CM, Tommet D, Schmitt EM, Travison TG, et al. The shortterm and long-term relationship between delirium and cognitive trajectory in older surgical patients. Alzheimers Dement. 2016;12(7):766–75.

10. Steinmetz J, Christensen KB, Lund T, Lohse N, Rasmussen LS, Group I. Long-term consequences of postoperative cognitive dysfunction. Anesthesiology. 2009;110(3):548–55.

11. Yurek F, Zimmermann JD, Weidner E, Hauss A, Dahnert E, Hadzidiakos D, et al. Quality contract ’prevention of postoperative delirium in the care of elderly patients’ study protocol: a non-randomised, pre-post, monocentric, prospective trial. BMJ Open. 2023;13(3):e066709.

12. Aldecoa C, Bettelli G, Bilotta F, Sanders RD, Audisio R, Borozdina A, et al. European Society of Anaesthesiology evidence-based and consensus-based guideline on postoperative delirium. Eur J Anaesthesiol. 2017;34(4):192–214.

13. Burton JK, Craig L, Yong SQ, Siddiqi N, Teale EA, Woodhouse R, et al. Non-pharmacological interventions for preventing delirium in hospitalised non-ICU patients. Cochrane Database Syst Rev. 2021;11(11):CD013307.

14. Lee S, Yu Y, Trimpert J, Benthani F, Mairhofer M, Richter-Pechanska P, et al. Virus-induced senescence is a driver and therapeutic target in COVID-19. Nature. 2021;599(7884):283–9.

15. Burry L, Mehta S, Perreault MM, Luxenberg JS, Siddiqi N, Hutton B, et al. Antipsychotics for treatment of delirium in hospitalised non-ICU patients. Cochrane Database Syst Rev. 2018;6(6):CD005594.

16. Miller D, Lewis SR, Pritchard MW, Schofield-Robinson OJ, Shelton CL, Alderson P, et al. Intravenous versus inhalational maintenance of anaesthesia for postoperative cognitive outcomes in elderly people undergoing non-cardiac surgery. Cochrane Database Syst Rev. 2018;8(8):CD012317.

17. Godbout JP, Johnson RW. Age and neuroinflammation: a lifetime of psychoneuroimmune consequences. Neurol Clin. 2006;24(3):521–38.

18. Aldecoa C, Bettelli G, Bilotta F, Sanders RD, Aceto P, Audisio R, et al. Update of the European Society of Anaesthesiology and Intensive Care Medicine evidence-based and consensus-based guideline on postoperative delirium in adult patients. Eur J Anaesthesiol. 2024;41(2):81–108.

19. Maldonado JR. Delirium pathophysiology: An updated hypothesis of the etiology of acute brain failure. Int J Geriatr Psychiatry. 2018;33(11):1428–57.

20. Dillon ST, Vasunilashorn SM, Otu HH, Ngo L, Fong T, Gu X, et al. Aptamer-Based Proteomics Measuring Preoperative Cerebrospinal Fluid Protein Alterations Associated with Postoperative Delirium. Biomolecules. 2023;13(9).

21. Egberts A, Mattace-Raso FU. Increased neutrophil-lymphocyte ratio in delirium: a pilot study. Clin Interv Aging. 2017;12:1115–21.

22. Li X, Cheng W, Zhang J, Li D, Wang F, Cui N. Early alteration of peripheral blood lymphocyte subsets as a risk factor for delirium in critically ill patients after cardiac surgery: A prospective observational study. Front Aging Neurosci. 2022;14:950188.

23. Lozano-Vicario L, Garcia-Hermoso A, Cedeno-Veloz BA, Fernandez-Irigoyen J, Santamaria E, Romero-Ortuno R, et al. Biomarkers of delirium risk in older adults: a systematic review and meta-analysis. Front Aging Neurosci. 2023;15:1174644.

24. Ko H, Kayumov M, Lee KS, Oh SG, Na KJ, Jeong IS. Immunological Analysis of Postoperative Delirium after Thoracic Aortic Surgery. J Chest Surg. 2024;57(3):263–71.

25. Lammers-Lietz F, Akyuz L, Feinkohl I, Lachmann C, Pischon T, Volk HD, et al. Interleukin 8 in postoperative delirium - Preliminary findings from two studies. Brain Behav Immun Health. 2022;20:100419.

26. Hall RJ, Watne LO, Idland AV, Raeder J, Frihagen F, MacLullich AM, et al. Cerebrospinal fluid levels of neopterin are elevated in delirium after hip fracture. J Neuroinflammation. 2016;13(1):170.

27. Osse RJ, Fekkes D, Tulen JH, Wierdsma AI, Bogers AJ, van der Mast RC, et al. High preoperative plasma neopterin predicts delirium after cardiac surgery in older adults. J Am Geriatr Soc. 2012;60(4):661–8.

28. Egberts A, Osse RJ, Fekkes D, Tulen JHM, van der Cammen TJM, Mattace-Raso FUS. Differences in potential biomarkers of delirium between acutely ill medical and elective cardiac surgery patients. Clin Interv Aging. 2019;14:271–81.

29. Kalantar K, LaHue SC, DeRisi JL, Sample HA, Contag CA, Josephson SA, et al. Whole-Genome mRNA Gene Expression Differs Between Patients With and Without Delirium. J Geriatr Psychiatry Neurol. 2018;31(4):203–10.

30. Song Y, Wang X, Hou A, Li H, Lou J, Liu Y, et al. Integrative Analysis of lncRNA and mRNA and Profiles in Postoperative Delirium Patients. Front Aging Neurosci. 2021;13:665935.

31. Saito T, Toda H, Duncan GN, Jellison SS, Yu T, Klisares MJ, et al. Epigenetics of neuroinflammation: Immune response, inflammatory response and cholinergic synaptic involvement evidenced by genome-wide DNA methylation analysis of delirious inpatients. J Psychiatr Res. 2020;129:61–5.

32. van Eijk MM, van Marum RJ, Klijn IA, de Wit N, Kesecioglu J, Slooter AJ. Comparison of delirium assessment tools in a mixed intensive care unit. Crit Care Med. 2009;37(6):1881–5.

33. Gaudreau JD, Gagnon P, Harel F, Tremblay A, Roy MA. Fast, systematic, and continuous delirium assessment in hospitalized patients: the nursing delirium screening scale. J Pain Symptom Manage. 2005;29(4):368–75.

34. Inouye SK, van Dyck CH, Alessi CA, Balkin S, Siegal AP, Horwitz RI. Clarifying confusion: the confusion assessment method. A new method for detection of delirium. Ann Intern Med. 1990;113(12):941–8.

35. Ely EW, Margolin R, Francis J, May L, Truman B, Dittus R, et al. Evaluation of delirium in critically ill patients: validation of the Confusion Assessment Method for the Intensive Care Unit (CAM-ICU). Crit Care Med. 2001;29(7):1370–9.

36. Kauffmann A, Gentleman R, Huber W. arrayQualityMetrics--a bioconductor package for quality assessment of microarray data. Bioinformatics. 2009;25(3):415–6.

37. Serpytis P, Navickas P, Navickas A, Serpytis R, Navickas G, Glaveckaite S. Age- and gender-related peculiarities of patients with delirium in the cardiac intensive care unit. Kardiol Pol. 2017;75(10):1041–50.

38. Galyfos GC, Geropapas GE, Sianou A, Sigala F, Filis K. Risk factors for postoperative delirium in patients undergoing vascular surgery. J Vasc Surg. 2017;66(3):937–46.

39. Shi C, Yang C, Gao R, Yuan W. Risk Factors for Delirium After Spinal Surgery: A Meta-Analysis. World Neurosurg. 2015;84(5):1466–72.

40. Oldroyd C, Scholz AFM, Hinchliffe RJ, McCarthy K, Hewitt J, Quinn TJ. A systematic review and meta-analysis of factors for delirium in vascular surgical patients. J Vasc Surg. 2017;66(4):1269–79 e9.

41. Vasunilashorn SM, Dillon ST, Marcantonio ER, Libermann TA. Application of Multiple Omics to Understand Postoperative Delirium Pathophysiology in Humans. Gerontology. 2023;69(12):1369–84.

42. Lammers-Lietz F, Akyuez L, Boraschi D, Borchers F, de Bresser J, Chatterjee S, et al. Development and internal validation of a gradient-boosted trees model for prediction of delirium after surgery and anesthesia (the BioCog study). medRxiv. 2024:2024.12.30.24319760.

43. Wang P, Wu Q, Shuai ZW. Emerging role of ficolins in autoimmune diseases. Pharmacol Res. 2021;163:105266.

44. Zhang J, Yang L, Ang Z, Yoong SL, Tran TT, Anand GS, et al. Secreted M-ficolin anchors onto monocyte transmembrane G protein-coupled receptor 43 and cross talks with plasma C-reactive protein to mediate immune signaling and regulate host defense. J Immunol. 2010;185(11):6899–910.

45. Ballweg T, White M, Parker M, Casey C, Bo A, Farahbakhsh Z, et al. Association between plasma tau and postoperative delirium incidence and severity: a prospective observational study. Br J Anaesth. 2021;126(2):458–66.

46. Khan SH, Lindroth H, Jawed Y, Wang S, Nasser J, Seyffert S, et al. Serum Biomarkers in Postoperative Delirium After Esophagectomy. Ann Thorac Surg. 2022;113(3):1000–7.

47. Spies CD, Knaak C, Mertens M, Brockhaus WR, Shadenok A, Wiebach J, et al. Physostigmine for prevention of postoperative delirium and long-term cognitive dysfunction in liver surgery: A double-blinded randomised controlled trial. Eur J Anaesthesiol. 2021;38(9):943–56.

48. MacLullich AM, Edelshain BT, Hall RJ, de Vries A, Howie SE, Pearson A, et al. Cerebrospinal fluid interleukin-8 levels are higher in people with hip fracture with perioperative delirium than in controls. J Am Geriatr Soc. 2011;59(6):1151–3.

49. van Munster BC, Zwinderman AH, de Rooij SE. Genetic variations in the interleukin-6 and interleukin-8 genes and the interleukin-6 receptor gene in delirium. Rejuvenation Res. 2011;14(4):425–8.

50. Lacy P. Mechanisms of degranulation in neutrophils. Allergy Asthma Clin Immunol. 2006;2(3):98–108.

51. Rorvig S, Honore C, Larsson LI, Ohlsson S, Pedersen CC, Jacobsen LC, et al. Ficolin-1 is present in a highly mobilizable subset of human neutrophil granules and associates with the cell surface after stimulation with fMLP. J Leukoc Biol. 2009;86(6):1439–49.

52. Hemmingsen R, Kramp P. Haematological changes and state of hydration during delirium tremens and related clinical states. Acta Psychiatr Scand. 1980;62(5):511–8.

53. Atkins JL, Pilling LC, Heales CJ, Savage S, Kuo CL, Kuchel GA, et al. Hemochromatosis Mutations, Brain Iron Imaging, and Dementia in the UK Biobank Cohort. J Alzheimers Dis. 2021;79(3):1203–11.

54. Lammers-Lietz F, Borchers F, Feinkohl I, Hetzer S, Kanar C, Konietschke F, et al. An exploratory research report on brain mineralization in postoperative delirium and cognitive decline. Eur J Neurosci. 2024;59(10):2646–64.

55. Yu Y, Zhu J, Mi LZ, Walz T, Sun H, Chen J, et al. Structural specializations of alpha(4)beta(7), an integrin that mediates rolling adhesion. J Cell Biol. 2012;196(1):131–46.

56. Blair P, Flaumenhaft R. Platelet alpha-granules: basic biology and clinical correlates. Blood Rev. 2009;23(4):177–89.

57. Nunez MT, Chana-Cuevas P. New Perspectives in Iron Chelation Therapy for the Treatment of Neurodegenerative Diseases. Pharmaceuticals (Basel). 2018;11(4).

58. Olave I, Wang W, Xue Y, Kuo A, Crabtree GR. Identification of a polymorphic, neuron-specific chromatin remodeling complex. Genes Dev. 2002;16(19):2509–17.

59. Sokpor G, Xie Y, Rosenbusch J, Tuoc T. Chromatin Remodeling BAF (SWI/SNF) Complexes in Neural Development and Disorders. Front Mol Neurosci. 2017;10:243.

60. Lamkanfi M, Kanneganti TD. Caspase-7: a protease involved in apoptosis and inflammation. Int J Biochem Cell Biol. 2010;42(1):21–4.

61. Shang Z, Lv H, Zhang M, Duan L, Wang S, Li J, et al. Genome-wide haplotype association study identify TNFRSF1A, CASP7, LRP1B, CDH1 and TG genes associated with Alzheimer’s disease in Caribbean Hispanic individuals. Oncotarget. 2015;6(40):42504–14.

62. Zhang X, Zhu C, Beecham G, Vardarajan BN, Ma Y, Lancour D, et al. A rare missense variant of CASP7 is associated with familial late-onset Alzheimer’s disease. Alzheimers Dement. 2019;15(3):441–52.

63. Sloan CD, Shen L, West JD, Wishart HA, Flashman LA, Rabin LA, et al. Genetic pathway-based hierarchical clustering analysis of older adults with cognitive complaints and amnestic mild cognitive impairment using clinical and neuroimaging phenotypes. Am J Med Genet B Neuropsychiatr Genet. 2010;153B(5):1060–9.

64. Burguillos MA, Deierborg T, Kavanagh E, Persson A, Hajji N, Garcia-Quintanilla A, et al. Caspase signalling controls microglia activation and neurotoxicity. Nature. 2011;472(7343):319–24.

65. van Gool WA, van de Beek D, Eikelenboom P. Systemic infection and delirium: when cytokines and acetylcholine collide. Lancet. 2010;375(9716):773–5.

66. Sun CC, Zhou ZQ, Yang D, Chen ZL, Zhou YY, Wen W, et al. Recent advances in studies of 15-PGDH as a key enzyme for the degradation of prostaglandins. Int Immunopharmacol. 2021;101(Pt B):108176.

67. Ribot JC, Lopes N, Silva-Santos B. gammadelta T cells in tissue physiology and surveillance. Nat Rev Immunol. 2021;21(4):221–32.

68. Feng L, Chen Y, Xu K, Li Y, Riaz F, Lu K, et al. Cholesterol-induced leucine aminopeptidase 3 (LAP3) upregulation inhibits cell autophagy in pathogenesis of NAFLD. Aging (Albany NY). 2022;14(7):3259–75.

