## supplement for "Altered gene expression associated with postoperative delirium in patients undergoing surgery and anesthesia"

**Supplementary table S1:** Patient characteristics for POD analysis (n =599) by sex. Characteristics are given as absolute and relative numbers, except for those marked ^a^, which are given as mean with standard deviation.

| Diagnosis | **POD**  **n=126** |  | **No POD**  **n=437** |  |
| --- | --- | --- | --- | --- |
| Sex | **Female**  n=60  (47.62 %) | **male**  n=66  (52.38 %) | **Female**  n=186  (42.56 %) | **male**  n=287  (65.68 %) |
| Peripheral surgery | 22 (36.70 %) | 22 (33.34 %) | 144 (77.42 %) | 167 |
| Age^a^ | 73.65 (4.69) | 74.58 (5.72) | 72.39 (5.07) | 71.87(4.92) |
| Duration of anesthesia^a^ | 389.17 (275.95) | 361.63 (235.50) | 199.80 (118.48) | 226.74 (199.64) |
| BMI^a^ | 28.80 (6.56) | 26.66 (4.10) | 27.66 (5.50) | 26.92 (4.03) |
| ASA_(status I-III) | 26 (43.34 %) | 31 (46.97 %) | 130 (69.89 %) | 201 (70.03 %) |
